# Understanding the clinical characteristics and timeliness of diagnosis for patients diagnosed with Long COVID: A retrospective observational cohort study from North West London

**DOI:** 10.1101/2024.08.30.24312849

**Authors:** Denys Prociuk, Jonathan Clarke, Nikki Smith, Ruairidh Milne, Cassie Lee, Simon de Lusignan, Ghazala Mir, Johannes De Kock, Erik Mayer, Brendan C Delaney, LOCOMOTION Consortium

## Abstract

**Background:** Long COVID is a multisystem condition first identified in the COVID-19 pandemic characterised by a wide range of symptoms including fatigue, breathlessness and cognitive impairment. Considerable disagreement exists in who is most at risk of developing Long COVID, driven in part by incomplete coding of a Long COVID diagnosis in medical records.

**Methods:** This was a retrospective observational cohort study using an integrated primary and secondary care dataset from North West London, covering over 2.7 million patients. Patients with Long COVID were identified through clinical terms in their primary care record. Multivariate logistic regression was used to identify factors associated with having Long COVID diagnosis, while multivariate quantile regression was used to identify factors predicting the time a Long COVID diagnosis was recorded.

**Findings:** A total of 6078 patients were identified with a Long COVID clinical term in their primary care record, 0.33% of the total registered adult population. Women, those aged 41 to 70 years or of Asian ethnicity were more likely to have a recorded Long COVID diagnosis, alongside those with pre-existing anxiety, asthma, depressive disorder or eczema and those living outside of the most socioeconomically deprived areas. Men, those aged 41 to 70 years, or of black ethnicity were diagnosed earlier in the pandemic, while those with depressive disorder were diagnosed later.

**Interpretation:** Long COVID is poorly coded in primary care records, and significant differences exist between patient groups in the likelihood of receiving a Long COVID diagnosis. Long COVID is more likely in those with pre-existing long-term conditions and is also associated with the frequent incidence of new long-term conditions. The experience of patients with Long COVID provides a crucial insight into inequities in access to timely care for complex multisystem conditions, and the importance of effective health informatics practices to provide robust, timely analytical support for front-line clinical services.

**Funding:** National Institute for Health and Care Research (NIHR) Ref: COV-LT2-0016

## Introduction

Over four years since the start of the coronavirus (COVID-19) pandemic, health systems are still uncertain on how and where best to diagnose and treat Long COVID, a complex multisystem condition resulting from acute COVID-19 infection (1). Current evidence suggests Long COVID may represent a diverse set of pathophysiological processes including viral persistence or reactivation, autoimmune responses, end organ damage and autonomic dysfunction (2,3). This in turn may produce a wide range of symptoms and clinical sequelae for patients, influenced by the particular way in which Long COVID has manifested, and how it interacts with an individual’s pre-existing conditions (1,4–7).

As a new condition without established clinical pathways, patients may experience challenges in navigating the healthcare system to obtain an appropriate and timely diagnosis. This is particularly the case for Long COVID where its symptom profile is particularly broad, encompassing a diverse range of symptoms including fatigue, breathlessness, cognitive dysfunction and palpitations (1,4).

There is evidence that electronic health records under-record the prevalence of Long COVID. Self-reported prevalence of Long COVID has been estimated at 3.3% in England and Scotland by the Office for National Statistics, between 6.6% and 10.4% in a national survey in Scotland and 7% in a large survey from the USA (8–10). Conversely, studies examining the presence of clinical codes for Long COVID in patient records estimate a prevalence of 0.02% (11). Others have found that of those with self-reported Long COVID, only 5.4% had a recorded diagnosis in their medical record, rising to 6.3% for those with severe symptoms, suggesting significant under-coding or under-diagnosis of Long COVID (12).

Several studies have identified factors associated with a Long COVID diagnosis. Across studies, women and those in middle and older age are more likely to self-report Long COVID symptoms and receive a Long COVID diagnosis (8,9,13,14). Similarly, vaccination against Covid-19 has been associated with reduced risk of Long COVID, while a diagnosis of one or more of several long term conditions including asthma, chronic obstructive pulmonary disease (COPD), anxiety, depression and diabetes have been associated with higher risk of developing Long COVID (8,9,13,14).

Considering ethnicity, white respondents were more likely to self-report Long COVID symptoms than black or Asian respondents to a US survey, and more likely to have a recorded Long COVID diagnosis in a UK study. Asian participants in the ONS Covid Infection Survey were, in contrast, more likely to report Long COVID symptoms (4.1%) than white (3.3%) and black participants (1.8%) (8,10,14). Adults living in regions in the most socioeconomically deprived quintile of England and Scotland were most likely to self-report Long COVID symptoms (5.6%) than those in the least deprived quintile (2.8%), but are less likely to have a recorded Long COVID diagnosis (10,14,15).

Although showing some agreement, these studies highlight important differences between self-reported and diagnosed Long COVID between patient groups. This may indicate greater barriers to receiving a Long COVID diagnosis for some patients than others. To examine this further, this study aims to determine, using an integrated electronic patient record dataset, the scale of Long COVID diagnosis in the population of North West London. We aim to examine factors that may be predictive of receiving a Long COVID diagnosis, the time of diagnosis, explore the clinical and demographic characteristics of these patients and quantify the prevalence of new long-term conditions amongst patients with Long COVID.

## Methods

### Data sources

The Whole Systems Integrated Care dataset (WSIC) is the primary source of data used for this study. WSIC is an integrated clinical database containing record level data derived from primary care records of over 2.7 million patients registered to participating GP practices in North West London.

### Data Analysis

Patients were identified as having Long COVID if their primary care record contained one of four Long COVID Systematized Nomenclature of Medicine (SNOMED) clinical terms after 1^st^ January 2020. These codes were chosen based on their suggested use in clinical coding by NHS England upon their creation as SNOMED codes in January 2021 (16). Other studies have used two UK-specific SNOMED clinical terms for Long COVID, specifically 1325161000000102 (Post-COVID-19 syndrome) and 1325181000000106 (Ongoing symptomatic COVID-19), however our study also includes international SNOMED codes to account for variation in coding norms between general practitioners, specifically 1119303003 (Post-acute COVID-19) and 1119304009 (Chronic post-COVID-19 syndrome) (16,17).

The presence of diagnosis clinical terms for each of 20 long term conditions (listed in Table 1) were identified using lists of SNOMED codes assigned to each condition obtained from the Oxford Royal College of General Practitioners Clinical Informatics Digital Hub (ORCHID) database (18). The date each condition first appears in a patient’s clinical record was also noted.

**Table 1.** Demographic and clinical characteristics of patients with a recorded Long Covid diagnosis and the overall adult WSIC patient population.

|  |  | Long Covid Patients |  | All Patients |  |
| --- | --- | --- | --- | --- | --- |
|  |  | N | % | N | % |
| Age | 18-30 | 788 | 12.96 | 280,425 | 15.25 |
|  | 31-40 | 1256 | 20.66 | 454,324 | 24.71 |
|  | 41-50 | 1392 | 22.90 | 381,394 | 20.75 |
|  | 51-60 | 1274 | 20.96 | 293,702 | 15.98 |
|  | 61-70 | 803 | 13.21 | 206,131 | 11.21 |
|  | 71-80 | 366 | 6.02 | 132,383 | 7.20 |
|  | 81-90 | 160 | 2.63 | 71,108 | 3.87 |
|  | >90 | 39 | 0.64 | 18,896 | 1.03 |
| IMD Quintile | 1 (most deprived) | 1142 | 18.79 | 360,633 | 19.62 |
|  | 2 | 1235 | 20.32 | 374,218 | 20.36 |
|  | 3 | 1270 | 20.90 | 372,633 | 20.27 |
|  | 4 | 1214 | 19.97 | 365,417 | 19.88 |
|  | 5 (least deprived) | 1070 | 17.60 | 360,766 | 19.62 |
|  | Missing | 147 | 2.42 | 4,696 | 0.26 |
| Gender | Female | 3992 | 65.68 | 885,981 | 48.19 |
|  | Male | 2086 | 34.32 | 952,326 | 51.80 |
|  | Other | 0 | 0.00 | 28 | 0.00 |
|  | Missing | 0 | 0.00 | 28 | 0.00 |
| Ethnicity | Asian or Asian British | 1825 | 30.03 | 471,249 | 25.63 |
|  | Black or Black British | 557 | 9.16 | 150,300 | 8.18 |
|  | Mixed | 238 | 3.92 | 54,310 | 2.95 |
|  | Other ethnic groups | 561 | 9.23 | 201,218 | 10.95 |
|  | White | 2860 | 47.05 | 884,728 | 48.13 |
|  | Missing | 37 | 0.61 | 76,558 | 4.16 |
| Total |  | 6078 |  | 1838363 |  |

The age, sex and ethnicity of the Long COVID population were described and compared to patients without a diagnosis of Long COVID, along with the distribution of Index of Multiple Deprivation ranks (a measure of socioeconomic status) of the Lower Layer Super Output Area of residence of those with and without a diagnosis of Long COVID. The prevalence of the 20 long-term conditions in the Long COVID patient population from 1^st^ January 2017 to 31^st^ December 2019 was compared to those without Long COVID in the WSIC dataset. Multivariate logistic regression including the panel of demographic covariates and comorbidities was used to identify factors predictive of receiving a diagnosis of Long COVID. Age was grouped into decades, with the exception of those aged 18 to 30 who were grouped together, and treated as a categorical variable with those aged 18 to 30 as the reference group.

For those patients with a recorded Long COVID diagnosis, the time between the 1^st^ of January 2020 and the date of Long COVID diagnosis was calculated. Multivariate median regression was used to identify factors predictive of patients receiving a Long COVID diagnosis later than others. Age was recoded as described above.

The incidence of new clinical comorbidities in the period from 1^st^ January 2020 to 5^th^ December 2023 in the Long COVID population was described and compared to those without a recorded Long COVID diagnosis. The 1^st^ of January 2020 was chosen in this case to identify all additional clinical comorbidities acquired since the start of the pandemic. As a supplementary analysis, K-Means clustering was used to group patients with a Long COVID diagnosis according to the clinical comorbidities they had been diagnosed with before 1^st^ January 2020 and separately the new conditions acquired on or after 1^st^ January 2020. Optimal configurations were determined using the elbow method. The relationship between these two configurations was compared using a Sankey diagram.

### Software

Data extraction was performed using Microsoft SQL Server Management Studio 2018, data processing and analysis was conducted using Python version 3.7.9 and the Pandas version 1.3.2 and numpy version 1.19.5 libraries. Statistical analysis was performed using Stata version 15. Clustering was performed using R version 4.3.2.

### Ethics statement

Ethical approval for this study was granted by the Yorkshire & The Humber - Bradford Leeds Research Ethics Committee (reference: 21/YH/0276) and from the Health Research Authority’s Confidentiality Advisory Group under reference (reference: 303623).

## Results

The WSIC dataset contained clinical records for 1,838,363 adult patients registered to General Practices within Northwest London as of 5^th^ December 2023. Of these patients, 6,078 (0.33%) were identified as having a Long COVID diagnosis code. Figure 1 shows the number of new diagnoses of Long COVID recorded each month since 1st January 2020. A peak in diagnosis frequency is seen in the first months of 2021 corresponding to the introduction of Long COVID SNOMED diagnosis codes. Over time, a gradual decline in diagnosis frequency is seen, alongside a change in coding practices. The initially dominant ‘Post-COVID-19 syndrome’ (1325161000000102) code is increasingly superseded by the other three codes, creating a mixed coding picture in the most recent diagnoses.

**Figure 1.**
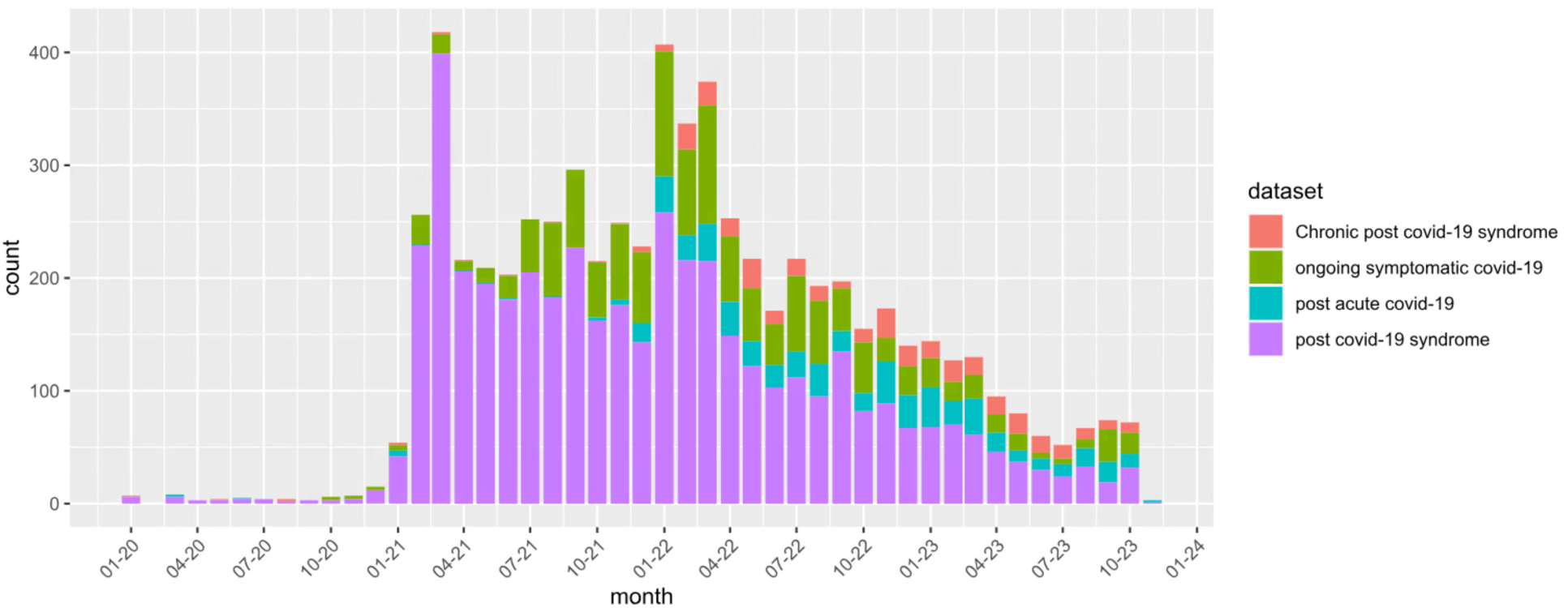
Frequency of new Long Covid diagnoses, disaggregated by the diagnosis code used.

Table 1 shows the demographic characteristics of the Long COVID population compared with the overall population of adult patients registered to GP practices within North West London. Patients with a Long COVID diagnosis were statistically significantly older, albeit with a small numerical difference (mean age 48.7 years vs. 48.0 years, p < 0.0001) and more likely to be female (65.7% vs. 48.2%, p < 0.0001) than those without a Long COVID diagnosis. Patients recorded as Asian or Asian British ethnicity accounted for 30.0% of Long COVID patients compared to 25.6% the overall population. Patients with missing ethnicity data represented 0.6% of the Long COVID population, but 4.2% of the overall WSIC population. Patients living in areas in the most socioeconomically deprived quintile of North West London accounted for 19.3% of those with a Long COVID diagnosis and 18.8% of those without a diagnosis while patients living in the least deprived quintile of areas accounted for 17.6% of patients with a Long COVID diagnosis and 19.6% of patients those without.

As shown in Table 2, before the COVID-19 pandemic, 2574 (42.3%) patients who went on to have a Long COVID diagnosis had one or more of the 20 included clinical comorbidities, compared to 3504 patients who did not. Compared with the overall population, asthma (12.9% vs. 3.8%), anxiety (12.4% vs. 3.3%), depressive disorder (11.0% vs. 3.3%), hypertension (10.6% vs. 4.7%), type 2 diabetes mellitus (7.7% vs. 3.4%) were more common in those with a recorded Long COVID diagnosis.

**Table 2.** Number of patients with a recorded diagnosis of each included clinical comorbidity in the period from January 2017 to 31st December 2019 compared to new conditions recorded after from 1st January 2020, excluding those with a pre-existing diagnosis of that condition. * denotes values below 5.

|  | 2017 - 2019 |  |  |  |  | 2020 2023 |  |  |  |  |
| --- | --- | --- | --- | --- | --- | --- | --- | --- | --- | --- |
|  | Long Covid Patients |  | All Patients |  | Chi-sq | Long Covid Patients |  | All Patients |  | Chi-sq |
| Condition | N | % | N | % | p | N | % | N | % | p |
| Anxiety | 752 | 12.37 | 84,049 | 4.57 | <0.001 | 982 | 18.44 | 71,210 | 4.06 | <0.001 |
| Asplenia | * | * | 272 | 0.01 | * | * | * | 296 | 0.02 | * |
| Asthma | 785 | 12.92 | 106,093 | 5.77 | <0.001 | 378 | 7.14 | 27,443 | 1.58 | <0.001 |
| Atrial Fibrillation | 67 | 1.10 | 24,266 | 1.32 | 0.1364 | 93 | 1.55 | 25,066 | 1.38 | 0.2711 |
| Chronic Kidney Disease | 91 | 1.50 | 20,120 | 1.09 | 0.0025 | 177 | 2.96 | 31,082 | 1.71 | <0.001 |
| Chronic Liver Disease | 5 | 0.08 | 2,372 | 0.13 | 0.309 | 15 | 0.25 | 2,369 | 0.13 | 0.0103 |
| Chronic Lung Disease | 75 | 1.23 | 17,753 | 0.97 | 0.0322 | 91 | 1.52 | 8,757 | 0.48 | <0.001 |
| Cirrhosis of liver | * | * | 1,391 | 0.08 | * | 10 | * | 1,533 | 0.08 | * |
| COPD | 78 | 1.28 | 17,834 | 0.97 | 0.0126 | 75 | 1.25 | 8,118 | 0.45 | <0.001 |
| Dementia | 22 | 0.36 | 8,846 | 0.48 | 0.1785 | 45 | 0.74 | 8,981 | 0.49 | 0.0049 |
| Depressive disorder | 668 | 10.99 | 87,399 | 4.75 | <0.001 | 811 | 14.99 | 62,020 | 3.54 | <0.001 |
| Diabetes Type 1 | 23 | 0.38 | 5,510 | 0.30 | 0.261 | 15 | 0.25 | 1,657 | 0.09 | <0.001 |
| Diabetes Type 2 | 468 | 7.70 | 105,875 | 5.76 | <0.001 | 261 | 4.65 | 39,766 | 2.30 | <0.001 |
| Eczema | 286 | 4.71 | 44,285 | 2.41 | <0.001 | 256 | 4.42 | 30,235 | 1.69 | <0.001 |
| Haemorrhagic stroke | 5 | 0.08 | 1,636 | 0.09 | 0.8601 | 10 | 0.16 | 2,284 | 0.12 | 0.3719 |
| Heart Failure | 30 | 0.49 | 10,804 | 0.59 | 0.3363 | 84 | 1.39 | 15,543 | 0.85 | <0.001 |
| Hypertension | 647 | 10.64 | 150,350 | 8.18 | <0.001 | 547 | 10.07 | 99,731 | 5.91 | <0.001 |
| Ischaemic Heart Disease | 77 | 1.27 | 19,182 | 1.04 | 0.086 | 130 | 2.17 | 18,628 | 1.02 | <0.001 |
| Ischaemic Stroke | * | * | 1,829 | 0.10 | * | 16 | * | 2,788 | 0.15 | * |
| Peripheral Vascular Disease | * | * | 970 | 0.05 | * | * | * | 1,502 | 0.08 | * |

### Factors predictive of receiving a Long COVID diagnosis

The above analyses reflect the univariate relationships between the clinical and demographic features of patents and their likelihood of receiving a Long COVID diagnosis. To account for correlation between these variables, multivariate logistic regression was performed to estimate the odds ratio for having a recorded Long COVID diagnosis accounting for a patient’s demographic and clinical characteristics before 1^st^ January 2020. Figure 2 shows the adjusted odds ratios for the multivariate logistic regression model for diagnosis of Long COVID.

**Figure 2.**
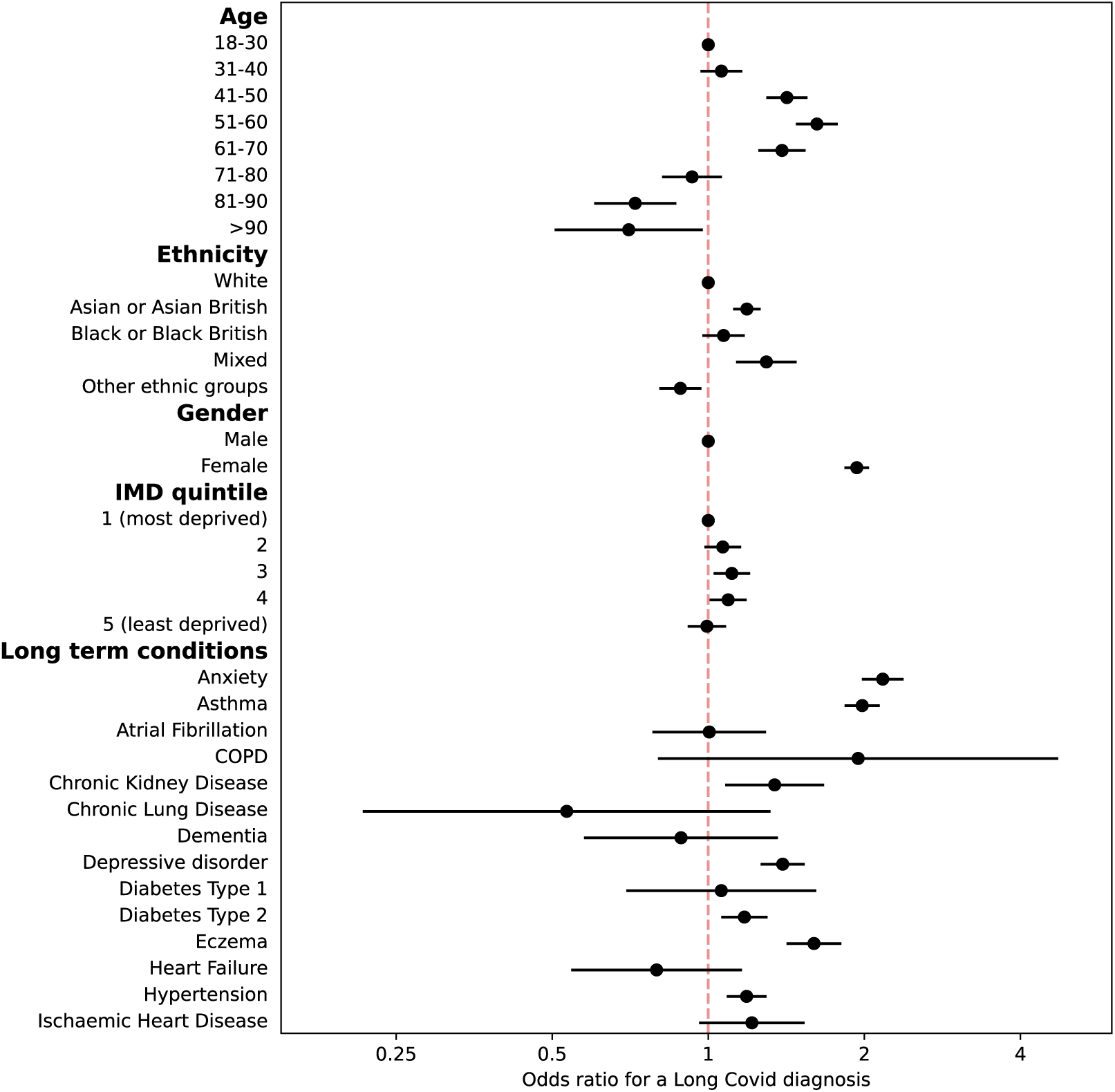
Adjusted odds ratios for having a recorded Long Covid diagnosis code in the primary care record. Comorbidities relate to conditions diagnosed before 1^st^ January 2020. Bars show 95% confidence intervals of the odds ratio.

Compared to the reference group of ages 18-30, those aged 31 to 70 years were significantly more likely to receive a Long COVID diagnosis, peaking at 51-60 years (aOR 1.62, p < 0.001). Those aged over 80 years were significantly less likely to receive a Long COVID diagnosis than those aged 18-30 years. Patients of Asian or Asian British, or mixed ethnicity were significantly more likely than patients of White ethnicity to receive a Long COVID diagnosis (aOR 1.19, and 1.29, p < 0.001). Patients whose ethnicity was recorded as ‘Other Ethnic Group’ had significantly lower odds of receiving a Long COVID diagnosis (aOR 0.88, p = 0.009).

Women were significantly more likely to have received a Long COVID diagnosis than men (aOR 1.93, p < 0.001). Those living in the least socioeconomically deprived quintile of areas were no more or less likely to have a recorded Long COVID diagnosis than those in the most deprived quintile. Those living in the third and fourth most deprived quintiles were significantly more likely to have a recorded Long COVID diagnosis than those in the most deprived quintile (aOR 1.11, p = 0.011 and 1.19, p = 0.035). Patients with recorded diagnoses of several long-term conditions before the Covid-19 pandemic were significantly more likely to receive a Long COVID diagnosis, specifically anxiety (aOR 2.17), asthma (aOR 1.98), eczema (aOR 1.59), chronic kidney disease (aOR 1.34), depressive disorder (aOR 1.39), hypertension (aOR 1.19) and type 2 diabetes (aOR 1.17). No conditions were associated with significantly lower odds of diagnosis of Long COVID, however small sample sizes for some less common conditions lead to uncertain estimates.

### Factors predictive of an earlier Long COVID diagnosis

Figure 3 shows the results of a multivariate median regression model estimating the time from the 1st of January 2020 to the date of Long COVID diagnosis for patients with a recorded Long COVID diagnosis. Compared to those aged 18-30 years, patients aged 41-70 years of age were diagnosed with Long COVID significantly earlier in the pandemic, with those aged 51-60 years estimated to be diagnosed an average of 62.9 days earlier than those aged 18-30. Compared to White patients, people recorded as Asian (-52.0 days, p < 0.001) or black ethnicity (-41.3 days, p = 0.01) were diagnosed significantly earlier in the pandemic. Women waited significantly longer than men (22.7 days, p = 0.015). No significant differences with respect to socioeconomic status were observed. Across the 20 conditions, only depressive disorder (32.7 days, p = 0.040) was associated with a statistically significant difference in the time of diagnoses, with large uncertainty in the estimates of less common conditions.

**Figure 3.**
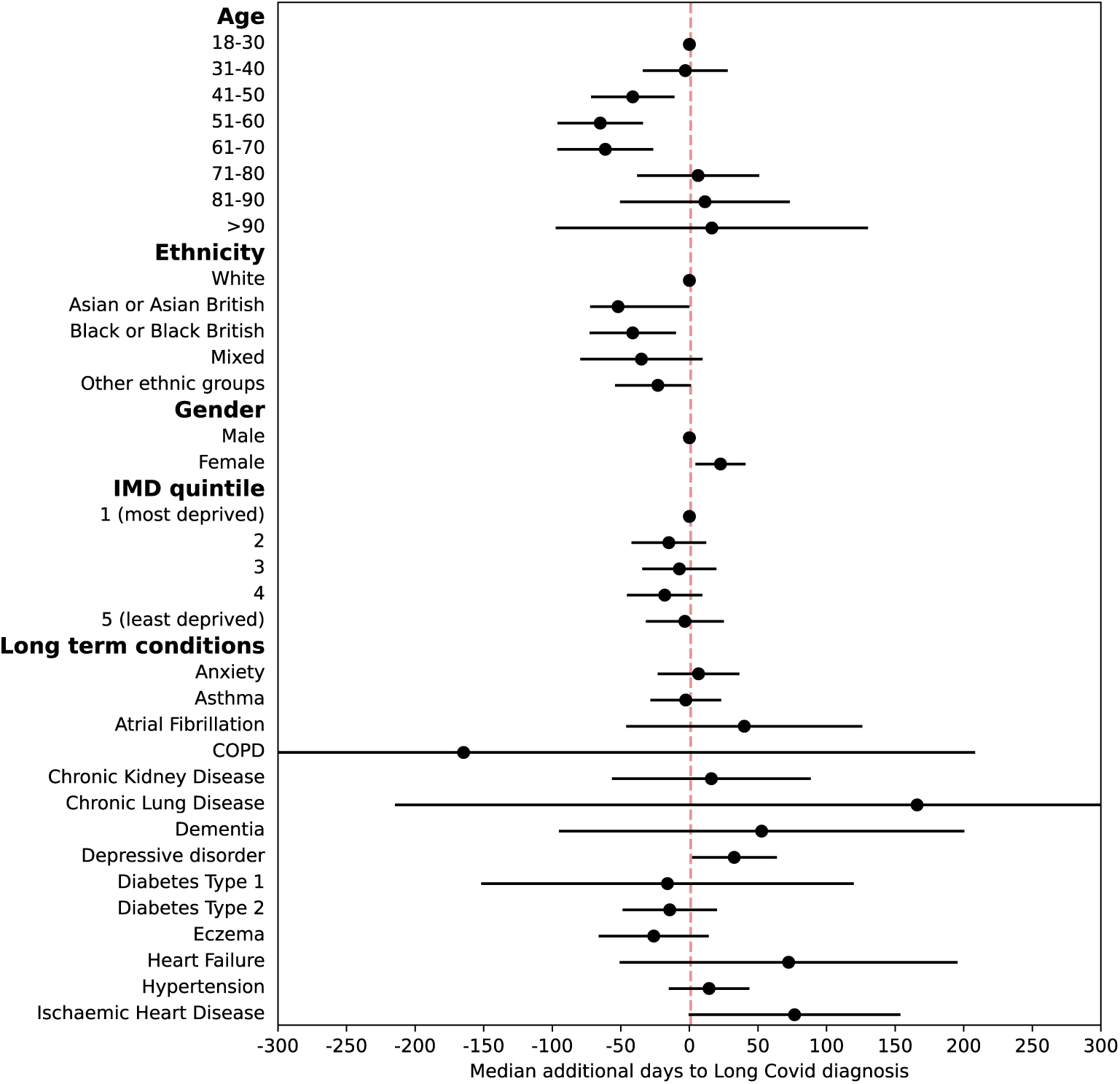
Predicted difference in median time to Long Covid diagnosis, obtained from a multivariate median quantile regression model. Comorbidities relate to conditions diagnosed before 1^st^ January 2020

### Acquisition of new conditions following a Long COVID diagnosis

Around 40% of patients (2574, 42.3%) with a diagnosis of Long COVID also had another new clinical condition entered into their primary care record in the period since the 1^st^ January 2020. As shown in Table 2, of those patients without a previous diagnosis of anxiety, 982 (18.4%) have received a new diagnosis of anxiety since 1st January 2020, compared to 4.1% of all patients. Similarly, of those without a previous diagnosis of depressive disorder, 811 (15.0%) have received a diagnosis of depressive disorder, compared to 3.5% overall. Other chronic conditions have also been newly diagnosed often, included hypertension (547 patients, 10.1%, vs. 5.9% overall), asthma (378 patients, 7.1%, vs. 1.6% overall) and Type 2 diabetes mellitus (261 patients, 4.7%, vs. 2.3% overall). As shown in the supplementary results, patients who go on to receive a Long COVID diagnosis may be grouped into distinct clusters based on their pre-existing conditions, if present. These clusters are largely centred around common conditions such as asthma, hypertension or Type 2 diabetes mellitus, with less marked contributions from a range of other diseases (Figure S2). Tracking these patients after the onset of the COVID-19 pandemic, we see a wide range of clinical trajectories in the development of new clinic comorbidities, with no dominant trajectories for patients based on their pre-existing conditions (Figure S5).

## Discussion

### Summary of findings

In this study of primary care record data from North West London, we find far lower prevalence of recorded Long COVID diagnoses (0.33%) than expected based on self-reported survey data from the UK Office for National Statistics (3.3%) and from a nationwide Scottish survey (6.6%- 10.3%), suggesting significant under-coding in primary care (9,10). This is supported by recent linkage of self-reported Long COVID to primary care records, finding as few as 5.4% of patients with self-reported Long COVID had a recorded Long COVID diagnosis (12). Using UK-specific and international SNOMED clinical concepts for Long COVID we see changes in coding practice over time, supporting the use of this wider set of codes in future analyses. Indeed, our Long COVID prevalence of 0.33% using diagnosis codes alone is far higher than the 0.02% prevalence observed using a similar method in Scotland (11).

Our finding of a recorded Long COVID diagnosis being more common in women and in those of middle age is consistently observed across other studies (9,11,14). We find those of Asian or mixed ethnicity were significantly more likely to receive a Long COVID diagnosis than white patients, a finding not observed in some studies, and the reverse identified in others (9,14,19). As in other studies, we find a trend towards higher risk of recorded Long COVID in those who do not live in the most socioeconomically deprived areas, however we do not find this association extends to the least deprived quintile of our sample (9,14). This aligns with a lower prevalence of Long COVID in the least deprived quintile of Scottish residents, and quartile of US residents (8,11).

Around four in ten patients (42.3%) who went on to receive a Long COVID diagnosis had a pre-existing long-term condition, with pre-existing asthma, eczema, anxiety and depressive disorder being strongly associated with a Long COVID diagnosis. Conversely, six in ten patients who received a Long COVID diagnosis had not been diagnosed with any of the twenty included clinical comorbidities before the pandemic. Since the onset of the COVID-19 pandemic however, patients who received a Long COVID diagnosis have experienced a significant increase in the burden of mental health conditions particularly, with 18.4% of those without a previous diagnosis of anxiety going on to receive a new diagnosis of anxiety since 2020, compared to 4.0% of the overall population. Although it is not possible to infer a causal link, similar patterns are seen for depressive disorder, suggesting an important role for mental health services in the management of Long COVID. Increased new diagnoses of hypertension, asthma and diabetes are observed, but again potential detection of common pre-existing conditions at a Long COVID-related consultation are quite likely. As the supplementary results indicate, patients with Long COVID have varied clinical trajectories in the new conditions they develop, rather than following a small set of patterns of disease.

An earlier diagnosis of Long COVID in the clinical record may reflect either earlier onset of Long COVID, a more timely recording of the diagnosis or a combination of the two. Thus, those exposed to COVID-19 earlier in the pandemic may be more likely to develop Long COVID earlier and therefore receive a recorded diagnosis earlier. We find middle-aged patients, those recorded as Asian, black or mixed ethnicity and men were more likely to be diagnosed with Long COVID earlier in the pandemic. This finding may partly be influenced by the disproportionate number of black and Asian workers in key sectors in London (20). Although 58% of key workers are female, large differences exist across occupations, with 90% of key workers in transport being male, while 79% of those in health and social care were female (21). Clinical comorbidities do not strongly influence the time of diagnosis, except for depression which was associated with diagnosis an average of 35 days later, suggesting potential barriers to receiving a timely diagnosis for those with poor mental health.

### Strengths and limitations

The study uses SNOMED codes from both the UK and International code lists to account for variation on Long COVID coding between practitioners. Using this strategy, we identify an additional 729 patients that would not otherwise have been included using UK codes alone. We also include codes for both Long COVID and Post-COVID conditions to identify patients with persistent symptoms after a COVID-19 infection.

Despite this strategy, it is likely that we have not identified a significant number of patients with Long COVID but for whom a relevant clinical term does not appear in their primary care record. This may result from diagnoses being made outside of primary care and not reported back to and/or coded by their general practitioner, patients who have not received a formal Long COVID diagnosis or where a code for another condition has been used.

We chose to focus on the relationship between Long COVID and a set of twenty common, relevant long-term conditions. These conditions represent a small subset of all long-term conditions, and it may be that associations with other conditions may emerge should a larger set be used. As this study includes only patients registered to GP practices in North West London, the findings of the study may not be generalisable to other regions. The region is entirely urban, and younger and more ethnically diverse than England as a whole.

In this study we were unable to precisely determine the date of COVID-19 infection and the date of onset of Long COVID symptoms. Instead, the 1^st^ of January 2020 was chosen over the date of a positive COVID-19 test as the index date for the time to diagnosis and to distinguish between pre- and post-pandemic clinic activity, for three reasons. Firstly, many patients within the dataset had multiple positive COVID-19 tests, separated over time and occurring both before and after their Long COVID diagnosis date. As Long COVID may result from the first or a subsequent COVID-19 infection, it is not possible to determine which positive COVID-19 test should be chosen. Secondly, many patients did not have a positive COVID-19 test in their clinical record. In these cases, using a COVID-19 test date to determine time to diagnosis would exclude these patients. Thirdly, feedback from the study’s Patient Advisory Group suggested that, particularly early in the COVID-19 pandemic, COVID-19 tests were not widely available and a diagnosis of COVID-19 was often based on clinical grounds alone, after which Long COVID symptoms developed. It was therefore not possible to precisely quantify the time from COVID-19 infection to the development of Long COVID symptoms or the diagnosis of Long COVID. Some conditions diagnosed after 1st January 2020 may predate COVID-19 infection or the occurrence of Long COVID symptoms, however taking the date of Long COVID diagnosis instead may exclude comorbidities diagnosed during the period between the onset of Long COVID symptoms and its diagnosis.

### Implications for policy and practice

The analysis of large health record datasets offers an important insight into how care is provided within a health system and how provision may vary between patient groups. Such studies rely on having a means to robustly identify the condition of interest, which in the case of Long COVID is notably lacking. A recent study used a range of methods to attempt to identify patients with Long COVID from primary care data, finding variable prevalence from 0.02% using diagnosis codes alone, 0.2% when using natural language processing to review clinical free text, 0.3% using data from sick notes and 1.7% when using a phenotype of clinical symptoms and patterns of activity (11). Crucially, the overlap between these different means of identification was generally low, supporting the use of multiple contrasting identification strategies.

Our study has demonstrated that delays in the introduction and implementation of Long COVID clinical terms precluded the timely recording of Long COVID diagnoses in health records. Specific clinical terms for Long COVID did not enter widespread clinical use until almost a year after the onset of the COVID-19 pandemic in the UK. Further, disagreement between NHS and international coding recommendations appear to have produced inconsistent coding practices between clinicians and over time (5,22,23). Consequently, incomplete, delayed or biased recording of a Long COVID diagnosis may in turn influence the findings of observational studies.

Although not all patients self-reporting Long COVID symptoms may meet the diagnostic criteria for the condition, the finding of at least a ten-fold difference in prevalence between the lower bound of national surveys (3.3%) and our study (0.33%) is deeply concerning. Efforts should continue to be made in primary and secondary care settings to identify and properly code patients who meet the diagnostic criteria for Long COVID.

While we find agreement between factors predisposing self-reported and diagnosed Long COVID for women, those in middle age and of Asian and white ethnicity, concern exists that those in the most socioeconomically deprived areas of North West London may be more likely to have Long COVID symptoms, but are not receiving a diagnosis at the same rate as those living in less deprived areas. Further work is also needed to understand these discrepancies and to address any associated unmet clinical need.

A coordinated approach to the clinical management of Long COVID is required to address both its wide-ranging symptoms, but also the high frequency with which patients with Long COVID go on to develop other long-term conditions, including mental health diagnoses.

While over four years have passed since the start of the Covid-19 pandemic in the UK, many patients continue to live each day with Long COVID. Quite how many is difficult to say, given delayed and incomplete diagnosis coding in clinical records, and much of what could have been learned about this new condition is to some extent now unknowable. Considering future pandemics, health systems must be more vigilant for the long-term sequelae of novel pathogens and rapidly develop appropriate clinical pathways to diagnose and treat patients. As part of this strategy, clinical terms must be developed and consistently implemented in tandem with these services to ensure health record data can be used to rapidly support the effective delivery of care.

## Conclusion

Long COVID occurs commonly within the UK population yet is significantly under-recorded in primary care records. The experience of patients with Long COVID provides a crucial insight into inequities in access to timely care for complex multisystem conditions, and the importance of effective health informatics practices to support robust and timely analytical support for front-line clinical services.

## Supporting information

LOCOMOTION consortium members

## Acknowledgements

The authors gratefully acknowledge infrastructure support from the National Institute for Health and Care Research (NIHR) Imperial Patient Safety Translational Research Centre, the NIHR Imperial Biomedical Research Centre and the NIHR Oxford Biomedical Research Centre. This research was in part enabled by the Imperial Clinical Analytics Research and Evaluation (iCARE) environment and Whole System Integrated Care and used the iCARE and WSIC team and data resources.

## Data availability statement

Data used for this study are available through application to DiscoverNOW (https://discover-now.co.uk/). Code used for the study and aggregate data outputs are available from the authors on reasonable request.

## Supplementary results

### Comorbidity clusters of Long COVID patients before and after the 1^st^ January 2020

K-means clustering was performed to identify Long COVID patients with similar profiles of pre-existing clinical comorbidities. 1,449 patients did not have a recorded clinical comorbidity prior to 1st January 2020 and were excluded, leaving 2474 patients included in the analysis. The elbow plot in Figure S1 identified a configuration consisting of 6 clusters to be optimal after 100 repeated initialisations to find the optimal initialisation values for each cluster centroid based on minimisation of the sum of squared distances from the centroid. Figure S2 shows the prevalence of each clinical comorbidity within each of the 6 clusters identified. The same process was repeated for conditions recorded on or after 1st January 2020. The elbow plot in Figure S3 identifies a configuration consisting of 9 clusters to be optimal and the frequency of each condition within a cluster is shown in Figure S4.

In the pre-pandemic period, four of six clusters are largely defined by the presence of a single comorbidity, namely anxiety, type-2 diabetes, hypertension and asthma (Figure S2). A further cluster represents the co-existence of anxiety and depression, while cluster 4 is a mixed cluster, with several conditions of low prevalence, with eczema being the most common. In the post-pandemic clustering of patients, clusters that are qualitatively similar to each of the clusters found in the pre-pandemic clustering are present, with three additional clusters identified (Figure S4). Two of these clusters are characterised by the coexistence of two conditions, specifically COPD and chronic lung disease (cluster 2) and type-2 diabetes and hypertension (cluster 6), while a cluster dominated by depressive disorder also appears. Figure S5 shows patients move between a wide range of clusters from the pre-pandemic to post-pandemic periods. This indicates that the further comorbidities patients with Long COVID may develop are varied and do not follow a narrow set of clinical trajectories.

---

**Figure S1.**
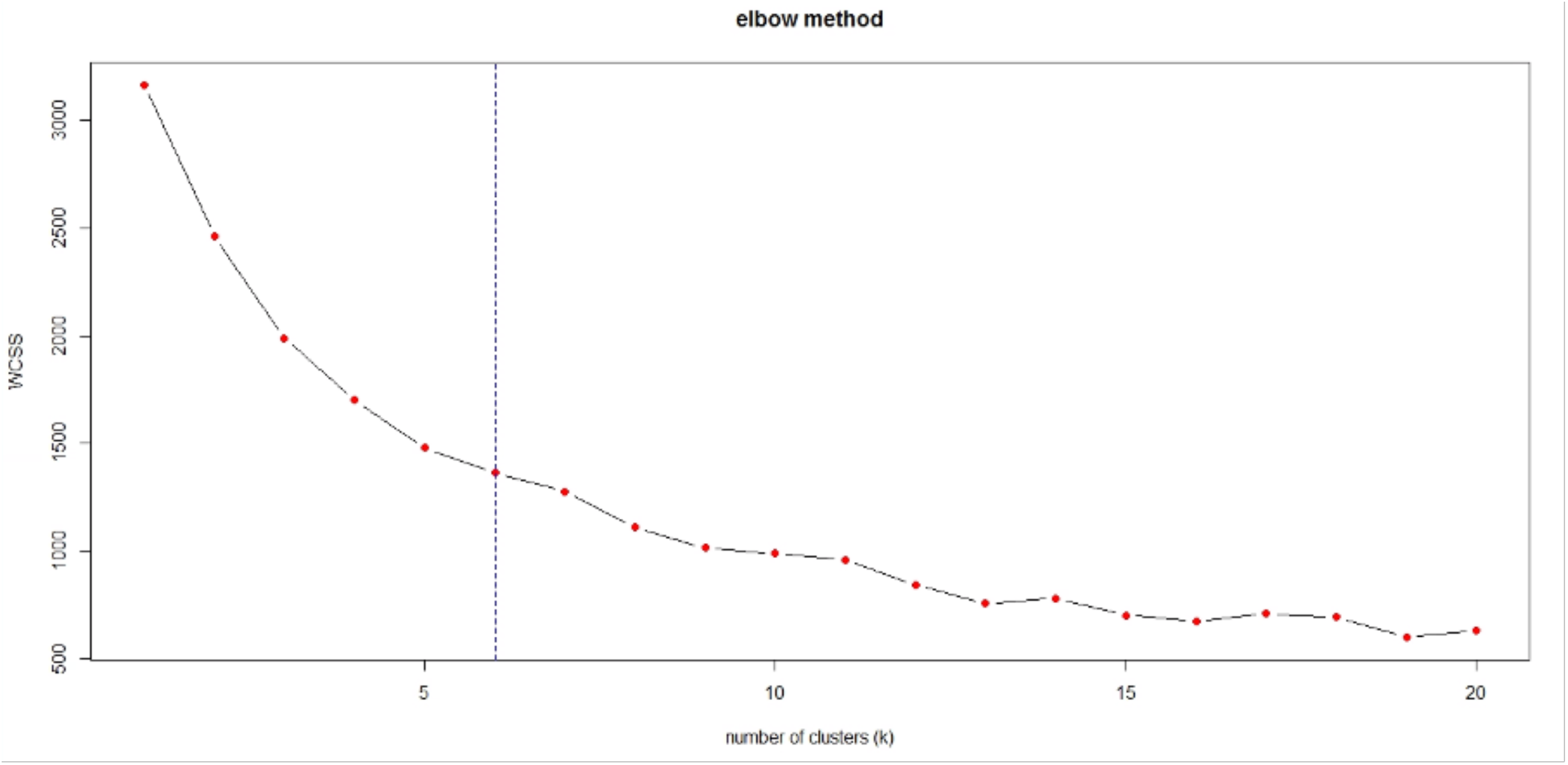
Elbow plot for K-means clustering of comorbidities for Long Covid patients between January 2017 and December 2019, vertical line indicates the plot chosen configuration of 6 clusters.

**Figure S2.**
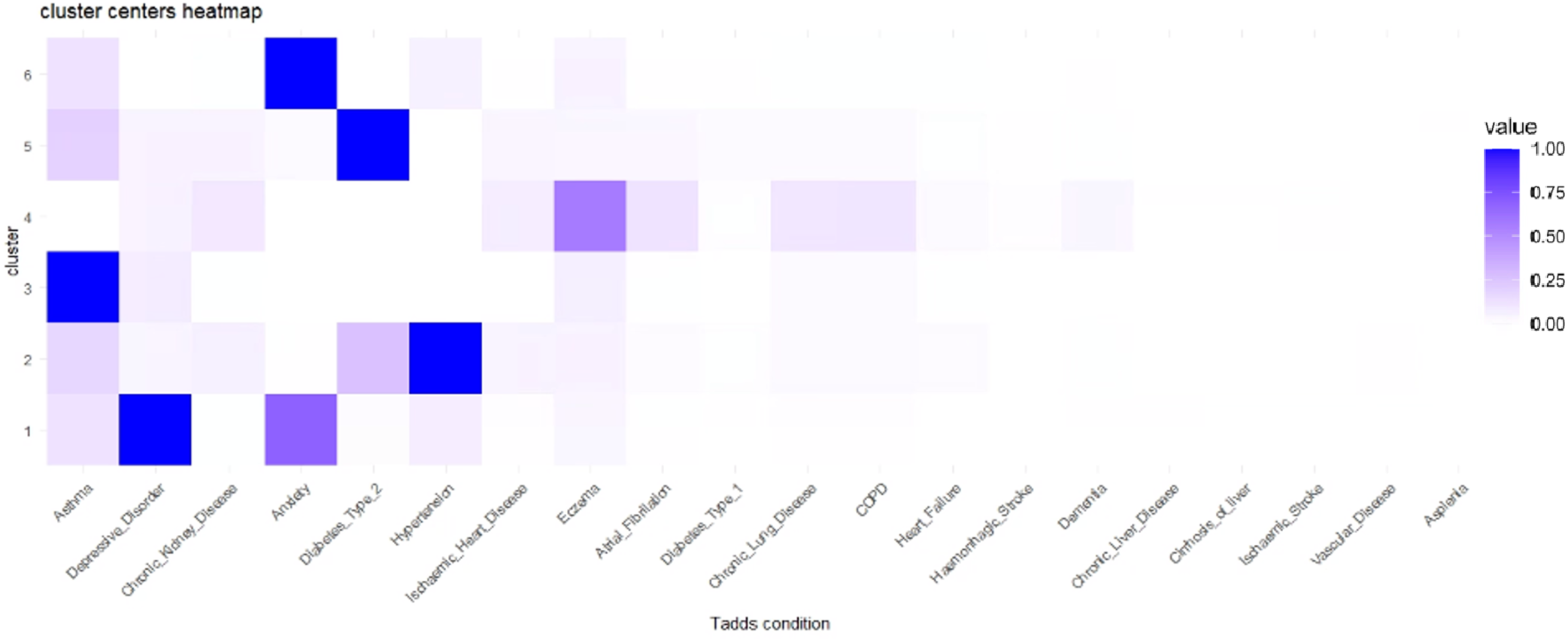
Heatmap showing the prevalence of conditions within each of the 6 patient clusters defined based on K-means clustering of comorbidities for Long Covid patients between January 2017 and December 2019.

**Figure S3.**
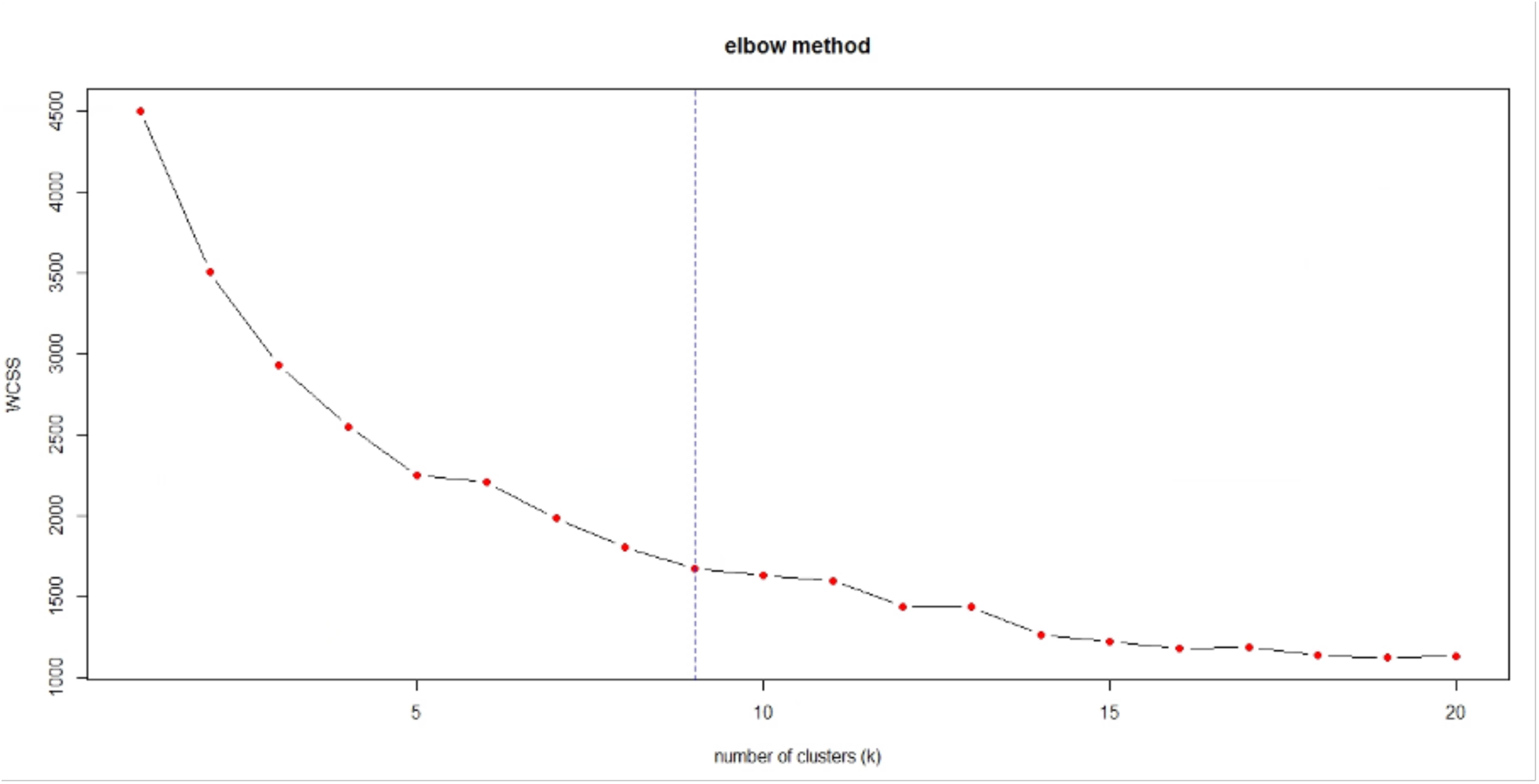
Elbow plot for K-means clustering of comorbidities for Long Covid patients between January 2020 and September 2023, vertical line indicates the plot chosen configuration of 9 clusters.

**Figure S4.**
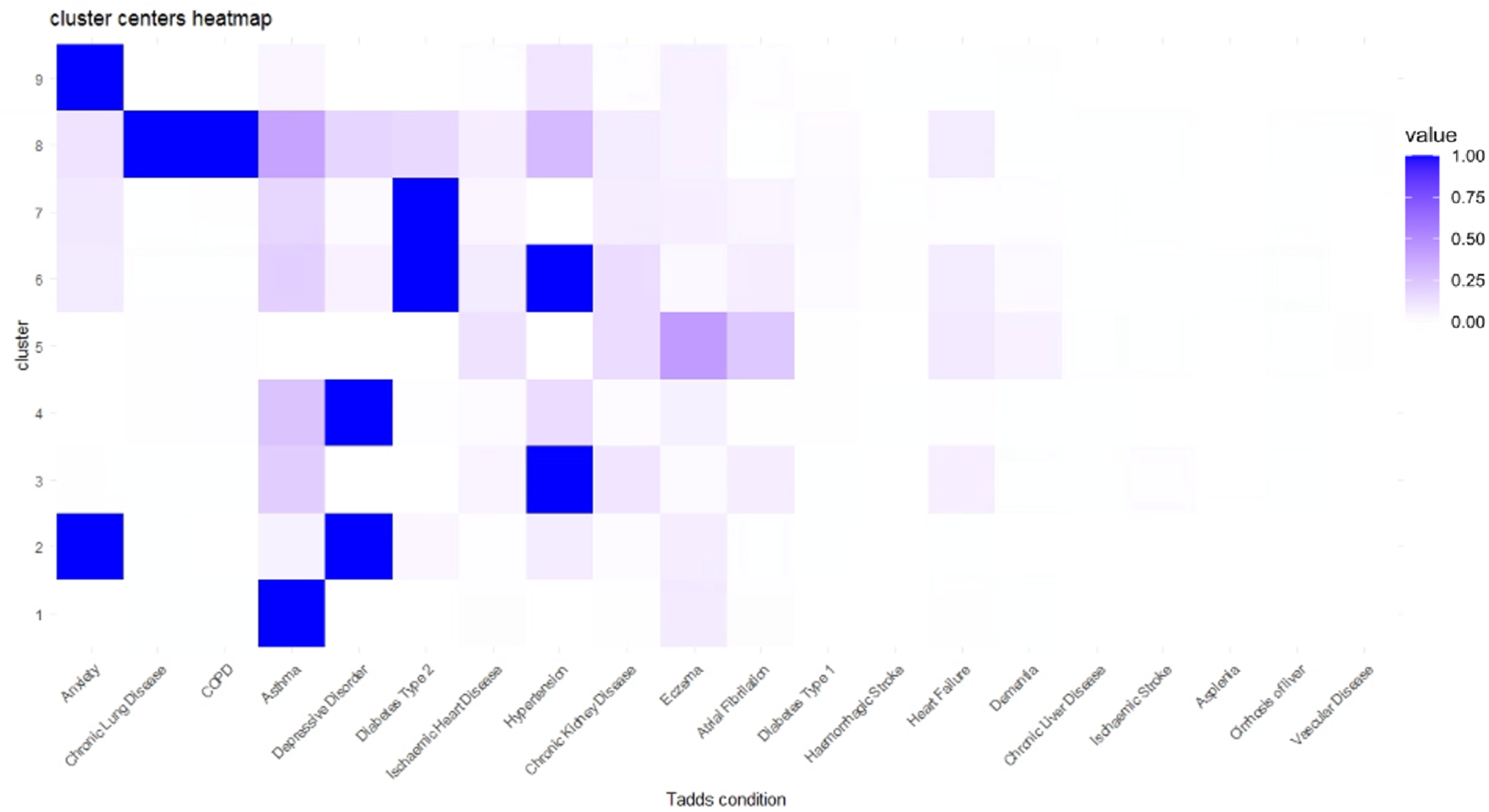
Heatmap showing the prevalence of conditions within each of the 9 patient clusters defined based on K-means clustering of comorbidities for Long Covid patients between January 2020 and September 2023.

**Figure S5.**
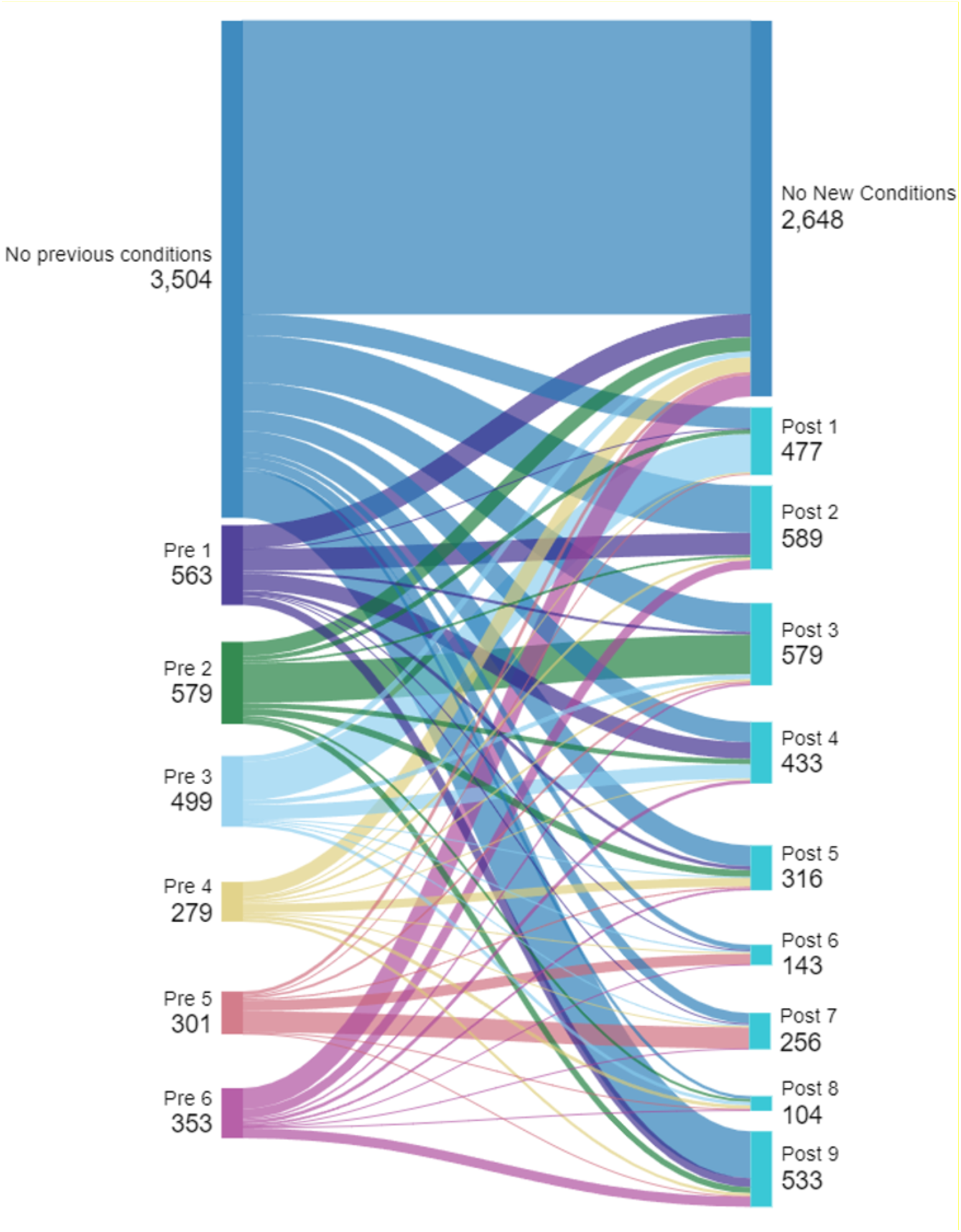
Sankey diagram showing the assignment of patients to disease clusters in the pre-pandemic and post-pandemic periods. Colours of edges represent the pre-pandemic cluster to which a patient was assigned. Numbers represent the total patients assigned to each cluster.

