## Supplementary material for "Understanding the clinical characteristics and timeliness of diagnosis for patients diagnosed with Long COVID: A retrospective observational cohort study from North West London": LOCOMOTION consortium members

**LOCOMOTION Consortium List**

| First name | Last name | Role |
|---|---|---|
| Nawar | Bakerly | Principal Investigator |
| Kumaran | Balasundaram | NHS Clinical Research Fellow |
| Megan | Ball | NHS Clinical Research Fellow |
| Mauricio | Barahona | Co-Investigator |
| Alexander | Casson | Co-Investigator |
| Jonathan | Clarke | HEI Researcher |
| Karen | Cook | Patient Advisory Group Member |
| Rowena | Cooper | NHS Clinical Research Fellow |
| Vasa | Curcin | Co-Investigator |
| Julie | Darbyshire | Co-Investigator |
| Helen | Davies | Principal Investigator |
| Helen | Dawes | Co-Investigator |
| Simon | de Lusignan | Co-Investigator |
| Brendan | Delaney | Chief Investigator |
| Carlos | Echevarria | Principal Investigator |
| Sarah | Elkin | Principal Investigator |
| Ana Belen | Espinosa Gonzalez | HEI Researcher |
| Rachael | Evans | Principal Investigator |
| Sophie | Evans | Patient Advisory Group Member |
| Zacchaeus | Falope | Principal Investigator |
| Ben | Glampson | HEI Researcher |
| Madeline | Goodwin | Research Assistant |
| Trish | Greenhalgh | Co-Investigator |
| Darren C | Greenwood | Co-Investigator |
| Stephen | Halpin | Principal Investigator |
| Juliet | Harris | NHS Research Assistant |
| Will | Hinton | HEI Researcher |
| Mike | Horton | Co-Investigator |
| Samantha | Jones | NHS Clinical Research Fellow |
| Joseph | Kwon | HEI Researcher |
| Cassie | Lee | NHS Clinical Research Fellow |
| Ashliegh | Lovett | NHS Clinical Research Fellow |
| Mae | Mansoubi | HEI Researcher |
| Victoria | Masey | NHS Clinical Research Fellow |
| Harsha | Master | Principal Investigator |
| Erik | Mayer | HEI Researcher |
| Bernardo | Meza-Torres | HEI Researcher |
| Ruairidh | Milne | Patient Advisory Group Member |
| Ghazala | Mir | Co-Investigator |
| Jacqui | Morris | Principal Investigator |
| Adam | Mosley | NHS Research Assistant |
| Jordan | Mullard | HEI Researcher |
| Daryl | O'Connor | Co-Investigator |

| | | |
|---|---|---|
| Rory | O'Connor | Co-Investigator |
| Thomas | Osborne | Project Manager |
| Amy | Parkin | NHS Clinical Research Fellow |
| Stavros | Petrou | Co-Investigator |
| Anton | Pick | Principal Investigator |
| Denys | Prociuk | HEI Researcher |
| Clare | Rayner | Patient Advisory Group Member |
| Amy | Rebane | Patient and Public Involvement Manager |
| Natalie | Rogers | Patient Advisory Group Member |
| Janet | Scott | Principal Investigator |
| Manoj | Sivan | Chief Investigator |
| Nikki | Smith | Patient Advisory Group Member |
| Adam | Smith | Statistician |
| Emma | Tucker | Principal Investigator |
| Ian | Tucker-Bell | Patient Advisory Group Member |
| Paul | Williams | NHS Clinical Research Fellow |
| Darren | Winch | Patient Advisory Group Member |
| Conor | Wood | NHS Research Assistant |
